# Plasma eMTBR-tau as a robust biomarker for Alzheimer’s disease tangle pathology up to 10 years before death

**DOI:** 10.64898/2026.09.29.26363951

**Authors:** Ruyu Shi, Xuemei Zeng, Marissa F. Farinas, Michel N. Nafash, Denis S. Smirnov, M. Ilyas Kamboh, Beth E. Snitz, Julia K. Kofler, Ann D. Cohen, Tharick A. Pascoal, Milos D. Ikonomovic, Henrik Zetterberg, Oscar L. Lopez, Thomas K. Karikari

**Affiliations:** Department of Psychiatry, School of Medicine, University of Pittsburgh, Pittsburgh, PA 15213, USA; Alzheimer’s Disease Research Center, University of Pittsburgh, Pittsburgh, PA, 15213, USA; Biofluid Biomarker Laboratory, Western Psychiatric Hospital, University of Pittsburgh Medical Center, Pittsburgh, PA, 15213, USA; Department of Pathology, Mass General Brigham, University of Pittsburgh, Pittsburgh, PA 15213, USA; Department of Genetics, School of Public Health, University of Pittsburgh, Pittsburgh, PA 15213, USA; Department of Pathology, School of Medicine, University of Pittsburgh, Pittsburgh, PA 15213, USA; Department of Neurology, School of Medicine, University of Pittsburgh, Pittsburgh, PA 15213, USA; Geriatric Research Education and Clinical Center, VA Pittsburgh HS, Pittsburgh, PA, USA; Department of Psychiatry and Neurochemistry, Institute of Neuroscience and Physiology, The Sahlgrenska Academy at the University of Gothenburg, S-431 80 Mölndal, Sweden; Clinical Neurochemistry Laboratory, Sahlgrenska University Hospital, S-431 80 Mölndal, Sweden; Department of Pathology and Laboratory Medicine, University of Wisconsin School of Medicine and Public Health, Madison, WI 53792, USA; Department of Neurodegenerative Disease, UCL Institute of Neurology, Queen Square, London WC1N 3BG, UK; UK Dementia Research Institute at UCL, London WC1N 3BG, UK; Centre for Brain Research, Indian Institute of Science, Bangalore, 560 012, India; Wisconsin Alzheimer’s Disease Research Center, University of Wisconsin School of Medicine and Public Health, University of Wisconsin-Madison, Madison, WI 53792, USA; Clinical and Translational Science Institute, School of Medicine, University of Pittsburgh, Pittsburgh, PA 15213, USA

## Abstract

Scalable and accessible biomarkers of neurofibrillary tangle pathology are needed for Alzheimer’s disease prognosis, diagnosis and monitoring. Here, we show that a blood-based immunoassay for an endogenously-cleaved “eMTBR-tau” fragment of tau protein is strongly associated with autopsy-diagnosed Alzheimer neuropathologic change (ADNC) and predominantly reflects the burden and anatomical progression of tangle pathology but not amyloid plaque load despite being most elevated in amyloid-positive symptomatic individuals. Plasma eMTBR-tau was significantly more accurate than plasma p-tau217 in classifying Braak staging and ADNC statuses at autopsy. Importantly, neuropathologic classification by plasma eMTBR-tau remained high (AUC=0.91-0.95) even when increasing intervals between blood collection and death up to a decade, while maintaining superior sensitivity and specificity over p-tau217. Higher baseline plasma eMTBR-tau was associated with worse cognition cross-sectionally and longitudinally predicted faster worsening. These results were replicated in a living cohort with *in vivo* tangle and plaque neuroimaging. Plasma eMTBR-tau concordance with tau neuroimaging was up to 94%, suggesting surrogacy. Together, plasma eMTBR-tau captures biological signatures of Braak staging and ADNC over a prolonged pre-mortem window, highlighting potential for early disease detection, biological staging, prognosis and therapeutic development.

## Main

The pathological hallmarks of Alzheimer’s disease (AD) – the progressive deposition of amyloid-beta (Aβ) plaques and neurofibrillary tangles (NFTs) composed of aggregated and hyperphosphorylated tau protein – are processes that begin years, potentially decades, before the onset and advancement of clinical symptoms.^1,2^ This window of opportunity provides an imperative for the development of high-performance biomarkers that can accurately detect and monitor these pathological changes during life.^1–3^

Blood-based biomarkers are transforming the diagnostic and prognostic evaluation of AD in living individuals via accessible and scalable means.^4^ However, the tests that are currently integrated into clinical and therapeutic trial programs mostly measure plasma phosphorylated tau 217 (p-tau217) which primarily detects Aβ pathology due to AD, as a “core 1” biomarker according to the revised Alzheimer’s Association diagnostic criteria.^5^ For this reason, AD-associated elevation of plasma p-tau217 often precedes the accumulation of NFTs, hence providing less resolution once NFT pathology is established.^6–8^ Scalable blood biomarkers that more directly track the extent of aggregated tau pathology – “core 2” biomarker – could therefore add clinically relevant information for biological staging, prognosis and trial selection targeting NFT-positive individuals.^9,10^

The microtubule-binding region of tau, as well as specific peptides within it, dictate the ability of the protein to aggregate into oligomers, paired helical filaments, and eventually NFTs.^11–15^ To this end, CSF assays that strongly correlate with NFT pathology all measure MTBR tau forms.^12,16,17^ In blood, an endogenous deamidated MTBR-containing tau fragment ending at residue 256 has emerged as one of the tau species most closely linked to AD NFT pathology, showing strong associations with cerebrospinal fluid MTBR-tau, tau PET burden, brain atrophy, and cognitive decline, with the greatest increases observed in symptomatic, amyloid- and tau-positive individuals.^9,10^ Studies combining mass-spectrometry-derived eMTBR-tau with p-tau217 further suggested that integrating these biomarkers may distinguish an upstream Aβ-associated phase from a later phase characterized by established NFT pathology.^18^ However, mass-spectrometry assays often require specialized and expensive equipment, large sample volumes and multiple pre-analytical processing steps, limiting scalability and ease of high-throughput deployment.^9,10^ Immunoassay alternatives for plasma eMTBR-tau that would address these concerns are urgently needed.

The relationship between plasma eMTBR-tau and AD neuropathology remains unknown as no previous study has investigated it. Here, we hypothesized that plasma eMTBR-tau would better reflect NFT pathology burden as well as AD neuropathological change (ADNC) status than plasma p-tau217 and that its diagnostic accuracy, specificity and sensitivity will remain high across an extended pre-mortem interval in the last decade of life. To test this, we evaluated the clinical performance of a novel low-volume, pre-processing-free immunoassay for plasma eMTBR-tau using Single-molecule proximity extension reaction (SPEAR) technology and an antibody highly sensitive for a defined region of the MTBR. We investigated the relationship between plasma eMTBR-tau and ADNC in a cohort of older adults with paired ante mortem plasma samples and subsequent neuropathological diagnosis at autopsy. These results were partly validated in an independent cohort with *in vivo* characterization Aβ plaque and NFT pathology using positron emission tomography (PET).

## Results

This study included two cohorts; one where participants had antemortem plasma sampling and neuropathologic characterization following death (the “autopsy cohort”) and another with plasma collection and neuroimaging assessments for Aβ and NFT abnormalities during life (the “PET cohort”). The autopsy cohort included 102 participants with antemortem SPEAR plasma eMTBR-tau measurements and postmortem neuropathologic assessment (**Extended Data Table 1**). Median age at blood draw was 75.5 years (interquartile range [IQR], 71.0-81.2 years), 49 participants (48.0%) were females and 75 (73.5%) had a clinical diagnosis of AD dementia. The median interval from blood draw to death was 4.9 years (IQR, 2.7-7.1). Braak stages V/VI and high ADNC were both present in 73 (71.6%) participants while 21 (20.6%) were classified as intermediate ADNC and 23 (22.5%) as Braak III/IV. The PET cohort included 137 participants (**Extended Data Table 2**); median age was 73.0 years (IQR, 68.0-79.0), 71 (51.8%) were female, 61 (44.5%) were Aβ-PET positive and 77 (56.2%) were tau-PET positive. Clinical diagnoses comprised 45 (32.8%) cognitively unimpaired controls, 42 (30.7%) participants with mild cognitive impairment (MCI), 35 (25.5%) with AD and 15 (10.9%) with other dementias.

In the autopsy cohort, plasma eMTBR-tau levels were higher with increasing disease severity according to each of the following metrics; clinical diagnosis, ADNC, Braak staging, Thal phases, and CERAD scores (**Fig. 1a; Supplementary Table 1a**). The median plasma eMTBR-tau level was 4.50-fold higher in participants with AD dementia vs. controls (p=0.003). Relative to Braak stages 0-II, the median plasma eMTBR-tau was 8.95-fold higher in Braak stages V/VI (p=9.80×10^-5^) individuals, and 8.08-fold higher in severe ADNC vs. low/not ADNC (p=1.20×10^-5^). These results were replicated in the PET cohort where plasma eMTBR-tau was higher according to clinical and Aβ-PET-defined severity (**Fig. 1b-c**). The median plasma eMTBR-tau was 4.47-fold higher in AD dementia vs. in controls (p=1.12×10^-9^) and 3.15-fold higher than in MCI (p=1.94×10^-6^) (**Fig. 1b, left**). When classified according to joint clinical and Aβ-PET status, plasma eMTBR-tau levels were highest in Aβ+ AD dementia followed in descending order by MCI Aβ+. The lowest levels were in CN Aβ-, CN Aβ+, MCI Aβ-and other (non-AD) dementias; these groups did not show any clear differences between them (**Fig. 1b, right; Supplementary Table 1b**). Together, these findings indicate that plasma eMTBR-tau levels are distinctively elevated in the AD continuum, with concentrations highest in those with advanced symptomatic disease (MCI Aβ+ and AD dementia).

**Fig. 1.**
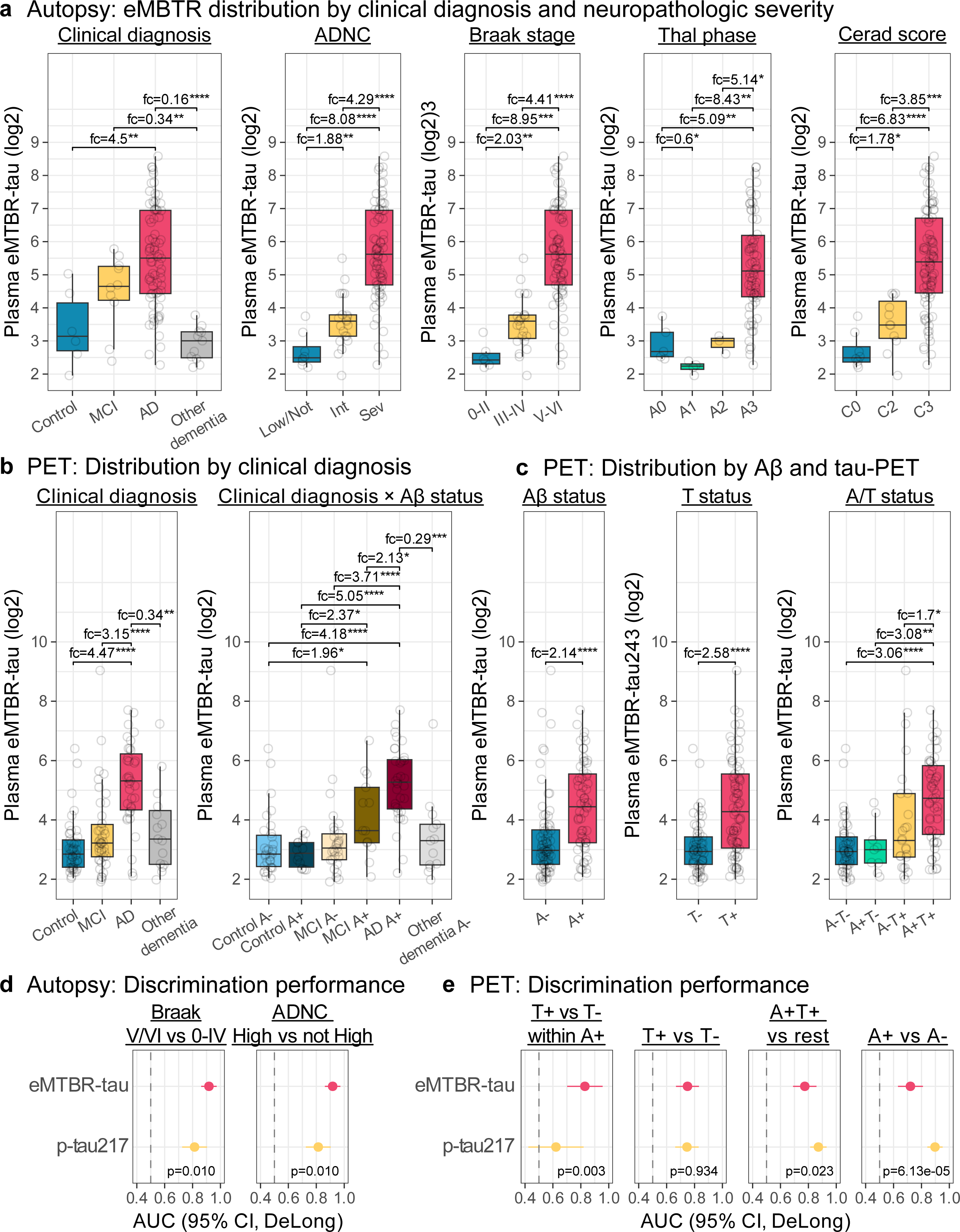
Plasma eMTBR-tau across autopsy and PET-defined Alzheimer disease severity and diagnostic performance. **a**, Plasma eMTBR-tau distributions in the autopsy cohort by clinical diagnosis, ADNC, Braak stage, Thal phase and CERAD neuritic plaque score. Brackets report significant pairwise median fold changes; points represent participants and boxes show the median and interquartile range. **b**, Plasma eMTBR-tau distributions in the PET cohort by clinical diagnosis and by clinical diagnosis stratified by amyloid-PET status. **c**, Plasma eMTBR-tau distributions in the PET cohort by amyloid-PET status, tau-PET status and combined imaging A/T status. **d**, Discrimination of Braak stages V/VI versus 0-IV and high versus not-high ADNC in the autopsy cohort by plasma eMTBR-tau and p-tau217. **e**, Discrimination in the PET cohort of tau-PET positivity among amyloid-PET-positive participants, tau-PET positivity in the full cohort, combined A+T+ status versus all other A/T groups and amyloid-PET positivity. Points and horizontal bars in **d** and **e** show AUC estimates and DeLong 95% confidence intervals; dashed vertical lines indicate an AUC of 0.5 and displayed p values are from paired DeLong tests. Fold-change comparisons used two-sided Wilcoxon tests with multiplicity-adjusted p values. Abbreviations: ADNC, AD neuropathologic change; A, amyloid; T, tau; PET, positron emission tomography; SUVR, standardized uptake value ratio; fc, fold change; AUC, area under the receiver operating characteristic curve.

Plasma eMTBR-tau was strongly associated with tau NFT pathology; the novel biomarker was more strongly associated with both ADNC (rho=0.67, p=2.32×10^-14^) and Braak staging (rho=0.76, p=1.81×10^-20^) than plasma p-tau217 (rho=0.50, p=1.76×10^-7^ for ADNC, bootstrap p=0.005 vs. plasma eMTBR-tau; and rho=0.55, p=3.13×10^-9^ Braak staging, bootstrap p=8.00×10^-4^ vs. plasma eMTBR-tau) in the autopsy cohort (**Extended Data Fig. 1a**). Corroborating results were obtained in the PET cohort, where plasma eMTBR-tau was numerically more strongly associated with tau-PET SUVR than plasma p-tau217, although the difference was not statistically significant (rho=0.60 versus 0.54; bootstrap p=0.34). In contrast, plasma p-tau217 was more strongly associated with amyloid-PET Centiloid than plasma eMTBR-tau (rho=0.67 versus 0.47; bootstrap p=0.004) (**Extended Data Fig. 1b**). Continuous analyses linked plasma eMTBR-tau more strongly to tau NFT than Aβ burden. In age- and sex-adjusted models, eMTBR-tau explained 44.3% of tau-PET SUVR variation, compared with 21.8% for Aβ-PET centiloid burden (both p≤1.51×10^-8^; **Extended Data Fig. 1c; Supplementary Table 2a**). Plasma eMTBR-tau was 2.14-fold and 2.58-fold higher in Aβ-PET and tau-PET positive vs. negative participants respectively (p=4.75×10^-6^ and p=2.52×10^-7^) (**Fig. 1c)**. According to combined Aβ and tau PET statuses, plasma eMTBR-tau was most elevated in the A+T+ group, being significantly higher than each of the A-T-, A-T+ and A+T-individuals (**Fig. 1c)**. Focusing on Aβ-PET positive participants who would qualify for anti-Aβ treatments provided they are symptomatic, plasma eMTBR-tau was 3.08-fold higher in tau-PET-positive than tau-PET-negative participants (p=0.005; **Supplementary Table 1b**). The results suggest that plasma eMTBR-tau is strongly associated with AD neuropathology and that it predominantly reflects the tau NFT burden component of ADNC than Aβ plaque load.

Next, we evaluated the capacity of plasma eMTBR-tau to distinguish individuals according to their ADNC and Braak staging statuses. In the autopsy cohort, Braak stage V/VI and high ADNC classifications were concordant (n=73), resulting in identical performance estimates. Plasma eMTBR-tau distinguished Braak stage V/VI from stages 0-IV and high ADNC from low and intermediate ADNC combined with an AUC of 0.92 (95% CI, 0.87-0.98; **Fig. 1d**; **Supplementary Table 3a**). For both outcomes, plasma eMTBR-tau outperformed plasma p-tau217 by an AUC difference of 0.11 (DeLong p=0.010; **Supplementary Table 3b**). In the PET cohort, plasma eMTBR-tau was superior to plasma p-tau217 in classifying tau-PET positivity with an AUC of 0.83 (95% CI, 0.70-0.96) vs. 0.62 (95% CI, 0.42-0.82) among Aβ-positive participants (Delong p = 0.003; **Fig. 1e**) but not in the full cohort (Delong p=0.934; **Fig. 1e; Supplementary Table 3c-d**). As expected, plasma p-tau217 was the better biomarker in identifying Aβ PET positivity (Delong p = 6.13×10^-5^; **Fig. 1e; Supplementary Table 3c-d**). The results suggest that plasma eMTBR-tau is more accurate than plasma p-tau217 for classifying the overall ADNC status at autopsy and captures the anatomical progression of tau NFT pathology whether determined by neuropathology or *in vivo* tau PET.

A central objective of this study was to determine how plasma eMTBR-tau measured years before death can accurately reflect ADNC and NFT Braak staging. Neuropathologic classification by plasma eMTBR-tau remained high even when increasing intervals between blood collection and death (**Fig. 2a** and **Extended Data Table 3**). Across 2-, 4-, 6-, 8- and 10-year windows, the AUCs to correctly identify Braak stages V/VI individuals remained high, being 0.95 (95%CI 0.84-1.00), 0.91 (95%CI 0.81-1.00), 0.92 (95%CI 0.85-0.99), 0.94 (95%CI 0.89-0.99) and 0.92 (95%CI 0.87-0.97), respectively. In the whole sample spanning the complete 10-year interval, sensitivity was 0.82 and specificity was 0.93 using the full-cohort Youden threshold. When compared with plasma p-tau217, plasma eMTBR-tau had significantly higher AUCs to detect Braak stages V/VI at 6, 8 and 10 years (AUC differences, 0.14, 0.14 and 0.11; Benjamini-Hochberg-adjusted p≤0.044 for each comparison). Additionally, plasma eMTBR-tau retained superior sensitivity and specificity at each time interval (**Fig. 2a** and **Extended Data Table 3**).

**Fig. 2.**
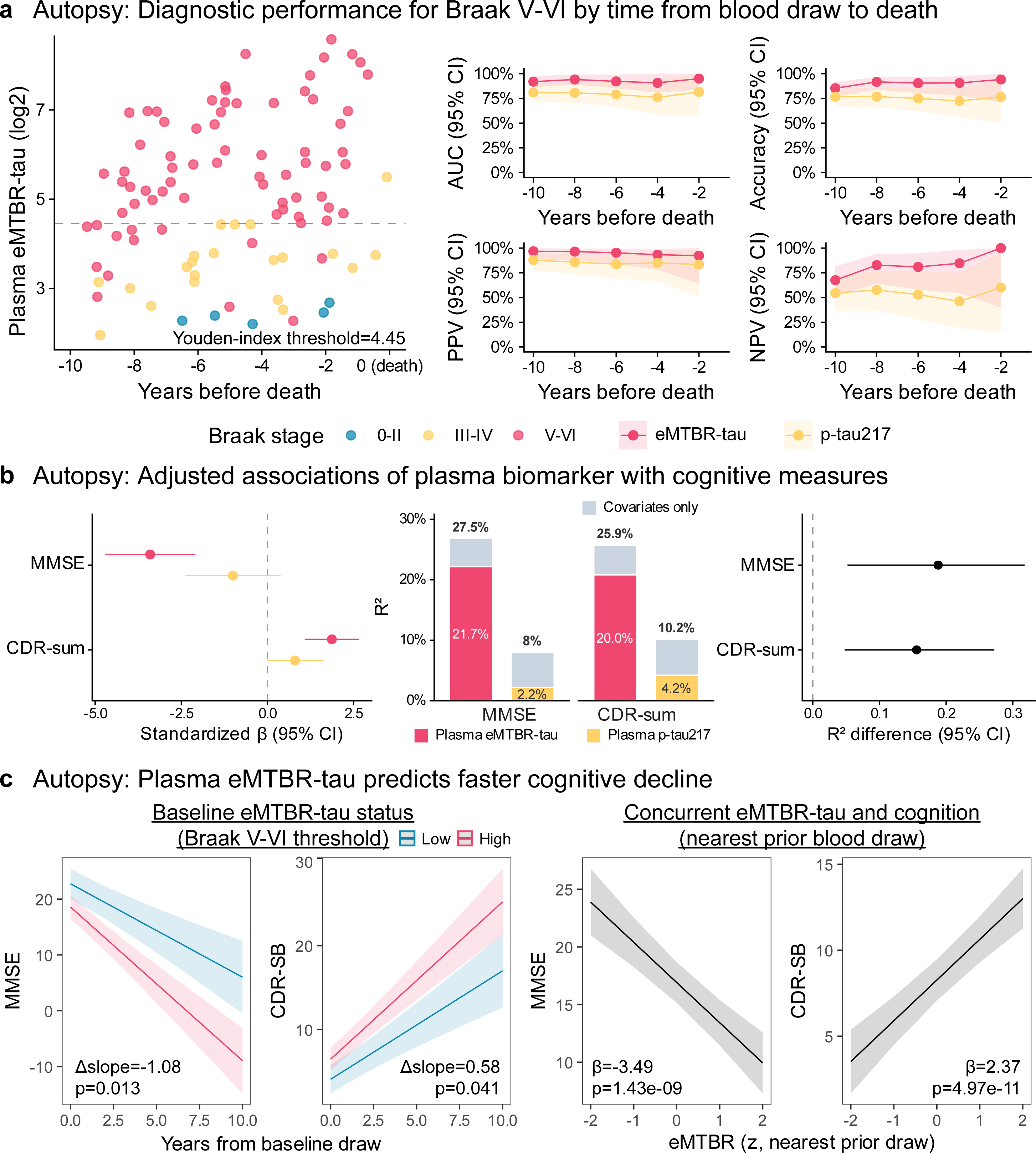
Temporal diagnostic performance and cognitive associations of plasma eMTBR-tau in the autopsy cohort. **a**, Plasma eMTBR-tau concentrations by years from blood draw to death, colored by Braak stage; the dashed horizontal line marks the full-cohort Youden threshold for Braak stages V/VI. Line plots show AUC, accuracy, positive predictive value (PPV) and negative predictive value (NPV) for plasma eMTBR-tau and p-tau217 across cumulative 2-, 4-, 6-, 8- and 10-year blood-draw-to-death windows; shaded bands denote 95% confidence intervals. **b**, Cross-sectional associations of plasma eMTBR-tau and p-tau217 with MMSE and CDR-Sum scores in the autopsy cohort. The left plot shows adjusted coefficients per 1-standard-deviation higher biomarker with 95% confidence intervals. Stacked bars partition model R2 into biomarker-attributable and covariate-only components, with total model R^2^ shown above each bar. The right plot shows paired-bootstrap differences in model R^2^ (eMTBR-tau minus p-tau217) with 95% confidence intervals. Models were adjusted for age, sex and education. **c**, Longitudinal cognitive analyses in the autopsy cohort. Left, model-predicted MMSE and CDR-Sum trajectories according to low versus high baseline plasma eMTBR-tau status defined by the Braak V/VI Youden threshold. Right, concurrent associations of standardized plasma eMTBR-tau from the nearest prior blood draw with MMSE and CDR-Sum. Lines show model-predicted values and shaded bands show 95% confidence intervals. Baseline-trajectory and concurrent mixed-effects models were adjusted for age, sex and education and included participant-specific random effects. Abbreviations: CI, confidence interval; CDR-Sum, CDR Sum of Boxes; MMSE, Mini-Mental State Examination.

Higher plasma eMTBR-tau at blood draw was associated with worse cognition in the autopsy cohort. For each 1-SD higher plasma eMTBR-tau, mini mental state examination (MMSE) was 3.34 points lower (n=101; 95% CI, −4.63 to −2.05; p=1.38×10^-6^), and CDR-Sum of Boxes (CDR-Sum) was 1.83 points higher (n=92; 95% CI, 1.05-2.61; p=1.13×10^-5^), after adjustment for age, sex and education (**Fig. 2b; Supplementary Table 4a**). Higher baseline plasma eMTBR-tau also predicted faster subsequent worsening, with plasma eMTBR-tau-positive participants showing faster worsening in MMSE (β=-1.08, p=0.013) and CDR-Sum (β=0.58, p=0.041) (**Fig. 2c left; Supplementary Table 4b**). Sensitivity analyses considering plasma eMTBR-tau as a continuous variable led to the same conclusion (**Supplementary Table 4c**). Supporting evidence was recorded in the PET cohort, where higher plasma eMTBR-tau was associated with worse MMSE performance (n=134; β=-1.16; 95% CI, −1.72 to −0.61; p=6.60×10^-5^) and higher CDR-Sum (n=134; β=1.10; 95% CI, 0.62-1.58; p=1.41×10^-5^), after adjustment for age, sex and education (**Extended Data Fig. 1d**).

In the autopsy cohort, plasma eMTBR-tau also tracked cognition over time. Matching each cognitive assessment to the nearest preceding blood draw, each 1-SD higher plasma eMTBR-tau was associated with 3.49 points lower MMSE (n=101, n observations=347; p=1.4×10^-9^) and 2.37 points higher CDR-Sum (n=92, n observations=338; p=5.0×10^-11^) (**Fig. 2c right**; **Supplementary Table 4d**). This association held both between individuals (MMSE β=-3.71, p=1.2×10^-7^; CDR-Sum β=2.16, p=2.4×10^-6^) and within individuals as plasma eMTBR-tau changed over time (MMSE β=-2.87, p=0.011; CDR-Sum β=2.76, p=6.8×10^-6^). In contrast, plasma p-tau217 showed only between-individual associations (MMSE β=-2.01, p=0.004; CDR-Sum β=1.01, p=0.027), with no within-individual association (MMSE p=0.95; CDR-Sum p=0.16) (**Supplementary Table 4e**). When both markers were modelled jointly, plasma eMTBR-tau was more strongly associated with concurrent cognition than plasma p-tau217 (contrast p=3.1×10^-9^ for MMSE; p=2.0×10^-12^ for CDR-Sum) (**Supplementary Table 4f**).

Next, we attempted to evaluate feasibility of using plasma eMTBR-tau as a surrogate measure of tau NFT pathology. When focusing on the Aβ-positive individuals alone, the Spearman correlation between plasma eMTBR-tau and tau PET was 0.73 (p=2.33×10^-11^**; Extended Data Fig. 2a**), with an overall accuracy of plasma eMTBR-tau to identify tau PET status at an encouraging 77.0% (**Extended Data Fig. 2a**). When separated into quadrants, the tau PET-positive/plasma eMTBR-tau-positive was the most concordant, accounting for 65.6% of all cases when the double negative quadrant accounted for 11.5%, indicating that nearly 77.0% of all cases were concordant. The overall concordance was improved to an impressive 93.9% when using a two-cutpoint approach that classified 12 (19.7%) cases in the intermediate zone (**Extended Data Fig. 2b**).

Finally, we investigated the viability of using plasma p-tau217 and plasma eMTBR-tau as surrogates for Aβ and tau-PET statuses. Single thresholds for plasma p-tau217 and eMTBR-tau was derived using the previously established cutpoints of 20 Centiloids for amyloid PET and an SUVR of 1.18 for tau PET (see Methods).^19^ Sequential plasma-based staging classified the four imaging-defined A/T groups with 56.3% (76/135) overall accuracy and 66.7% (90/135) accuracy for tau PET positivity alone (**Extended Data Fig. 2c**). When excluding the A-T+ cases who are considered individuals with suspected non-Alzheimer pathology^20^, the overall accuracy improved to 66.4% (73/110) while the tau PET accuracy raised to 75.5% (83/110). Notably, most A-T+ individuals (18/25) were classified as plasma eMTBR-tau-negative, suggesting that plasma eMTBR-tau reflects Alzheimer-type tau NFT pathology rather than tau PET signal arising from non-Alzheimer processes. Together, these results provide initial evidence that plasma eMTBR-tau could complement tau PET as a measure of tau NFT pathology.

## Discussion

We have demonstrated that plasma eMTBR-tau is strongly associated with – and accurately classifies – ADNC and tau NFT pathology, and more importantly, can identify individuals who will later be confirmed to have AD neuropathology at autopsy when blood is collected many years before their death. This association and high classification accuracies persisted over an interval extending to approximately a decade before death, providing defining evidence that circulating eMTBR-tau levels can capture the biological basis of tau NFT pathology over a prolonged period. These findings, externally validated in a cohort where NFT pathology was defined by *in vivo* tau PET measures where plasma eMTBR-tau was distinctively associated with and more strongly explained the variance in tau NFT pathology compared with Aβ plaque pathology, establish plasma eMTBR-tau as a unique blood-based measure of tau NFT pathology rather than a general marker of Aβ-associated AD biology.

The stability of these findings across years is particularly worth highlighting. Since AD pathophysiology starts many years before dementia onset and progresses biologically over a long continuum, biomarkers with limited resolution i.e., become abnormal only when pathology is advanced or when blood is collected as close as possible to death may have limited utility for contexts of use in pre-dementia stages such as prognosis, disease prevention campaigns and therapeutic evaluation and monitoring. To the contrary, a blood biomarker such as the SPEAR eMTBR-tau assay described herein capable of detecting and quantifying tau NFT burden and ADNC diagnosis years pre-death could provide meaningful way to prognose, diagnose and select older adults for inclusion in interventional programs.

Our results reinforce the importance of having accessible biomarkers with the capacity to distinguish between Aβ and tau NFT pathology, the principal pathological hallmarks of AD, as core 1 and core 2 biomarkers. As the MTBR sequence contains peptides that are crucial to the pathological tau aggregation cascade, the plasma eMTBR-tau immunoassay reports on brain tau aggregation status as revealed by the relationship between plasma eMTBR-tau and tau NFT pathology – determined by Braak staging at autopsy or *in vivo* tau PET.

The results have important implications for clinical diagnosis, eligibility for anti-Aβ treatments (AATs), disease prognosis as well as therapeutic trial and prevention program enrolment and monitoring. While plasma p-tau217 – the current leading core 1 blood biomarker with assays from multiple sources having received regulatory approval – accurately detects Aβ pathology due to AD^21^, it is suboptimal for classifying NFT pathology.^9,18,22^ Positivity for both plasma p-tau217 and eMTBR-tau increases the likelihood to identify the Aβ and tau NFT abnormalities that define AD diagnosis in accordance with current clinical practice guidelines.^9,18,23,24^ Concerning eligibility for AATs, therapeutic trial and prevention programs, relying solely on abnormal plasma p-tau217 levels is likely to inevitably select Aβ-positive individuals who may never progress to overt A+T+ or AD dementia profiles.^25^ Contrarily, selecting participants based on combined plasma eMTBR-tau and p-tau217 abnormalities may address this limitation.

The findings will also have utility for anti-AD therapeutic trials which continue to recruit individuals in early phases of the AD continuum, creating a need for scalable and cost-effective biomarkers for disease diagnosis and biological staging. A blood biomarker such as plasma eMTBR-tau that sensitively tracks tau NFT pathology in living individuals as well as years before death could facilitate screening, longitudinal monitoring and clinical trial recruitment, enabling repeated assessment of disease trajectories in settings where routine CSF measurement or PET imaging may not be feasible or preferred.

The results that solubilized or soluble tau assemblies that potentially act as seeds for tau aggregation and hence ongoing NFT accumulation over several to many years can be detected by the plasma eMTBR-tau assay point to an important implication: that this biomarker can potentially serve as a surrogate for tau PET in therapeutic and observational trials. This would help reduce the cost of clinical trial enrolment as well as expand access ad attractiveness of inclusion to communities that are reluctant to invasive procedures. Furthermore, as blood-soluble versions of the endogenously cleaved MTBR fragment are expected to become abnormal ahead of tau PET (and as the blood levels are predictive of future tangle load as shown here), plasma eMTBR-tau may be an excellent biomarker for monitoring disease progression as well as the efficacy of anti-tau treatments several of which are currently being evaluated in phases I-III clinical trials.

An important strength of the present study is the assessment of the direct link between antemortem blood biomarker measurements and neuropathology postmortem. Autopsy remains the most definitive means for establishing the neuropathological basis of AD, despite PET imaging and cerebrospinal fluid alternatives providing powerful measures. Thus, connecting plasma biomarker measures to subsequent neuropathological evaluation is an exceptionally rigorous framework for the clinical validation of new biomarker candidates.

This study has a few limitations. Firstly, the use of a selected autopsy cohort could bias inclusion for instance in terms of racial and ethnic identity. Thus, expansion of the findings to the broader population of individuals at risk for AD remains to be evaluated. Secondly, the variable intervals between blood collection and neuropathological assessments at death mean that plasma biomarker measures may not be contemporaneous with brain pathology. Finally, larger cohort studies with longitudinal plasma collection, more generalizable participant inclusion and external validation of cutpoints are needed.

In conclusion, the SPEAR plasma eMTBR-tau immunoassay is a novel blood-based measure of AD neuropathology based on its specific tracking of tau NFT pathology across Braak stages that remains stable for many years pre-death. Additionally, plasma eMTBR-tau demonstrated to be a durable measure of tau pathology that is detectable during older adulthood and showed stronger agreement with, and high classification accuracies for, *in vivo* NFT pathology than with Aβ plaque burden. These results establish plasma eMTBR-tau as a potential biomarker for tau NFT pathology, which warrants further evaluation for early detection, disease prognosis, biological staging as well as eligibility determination for AATs, therapeutic trial and prevention programs.

## Data Availability

Summary statistics, derived datasets, and analytic outputs sufficient to reproduce the main findings are available from the corresponding author upon request, subject to a data use agreement and approval by the University of Pittsburgh Alzheimer’s Disease Research Center.

## Acknowledgements

We thank the research participants and their families for their invaluable contributions. This study was supported by NIH/NIA grants P30 AG066468 (PI: Oscar L. Lopez), R01 AG083874, U24 AG082930, and U01 AG098641 (PI: Thomas K. Karikari). Dr. Karikari’s Biofluid Biomarker Laboratory, which houses the ADRC Biomarker Core, was further supported by NIH/NIA (RF1 AG077474, R01 AG083156, R37 AG023651, R01 AG025516, R01 AG073267, R01 AG075336, R01 AG072641, P01 AG025204, P01 AG14449, R01 AG013672, R01 AG041718, R01 AG030653, and R01 AG064877), NIH/NINDS (U01 NS131740 and U01 NS141777), NIH/NIMH (R01 MH108509), the Aging Mind Foundation (DAF2255207), the Department of Defense (HT94252320064), the Anbridge Charitable Fund, and a professorial endowment from the Department of Psychiatry, University of Pittsburgh. Dr. Zetterberg is a Wallenberg Scholar and a Distinguished Professor at the Swedish Research Council supported by grants from the Swedish Research Council (#2023-00356, #2022-01018 and #2019-02397), the European Union’s Horizon Europe research and innovation programme under grant agreement No 101053962, Swedish State Support for Clinical Research (#ALFGBG-71320), the Knut and Alice Wallenberg Foundation (KAW 2025.0225), the Alzheimer Drug Discovery Foundation (ADDF), USA (#201809-2016862), the AD Strategic Fund and the Alzheimer’s Association (#ADSF-21-831376-C, #ADSF-21-831381-C, #ADSF-21-831377-C, and #ADSF-24-1284328-C), the European Partnership on Metrology, co-financed from the European Union’s Horizon Europe Research and Innovation Programme and by the Participating States (NEuroBioStand, #22HLT07), the Bluefield Project, Cure Alzheimer’s Fund, the Olav Thon Foundation, the Erling-Persson Family Foundation, Familjen Rönströms Stiftelse, Familjen Beiglers Stiftelse, Stiftelsen för Gamla Tjänarinnor, Hjärnfonden, Sweden (#FO2022-0270), the European Union’s Horizon 2020 research and innovation programme under the Marie Skłodowska-Curie grant agreement No 860197 (MIRIADE), the European Union Joint Programme – Neurodegenerative Disease Research (JPND2021-00694), the National Institute for Health and Care Research University College London Hospitals Biomedical Research Centre, the UK Dementia Research Institute at UCL (UKDRI-1003), and an anonymous donor. The content of this article is solely the responsibility of the authors and does not necessarily represent the official views of the funders.

## Author Contributions Statement

Conception and experimental design: X.Z. and T.K.K. Cohort recruitment and retention: B.E.S., A.D.C., and O.L.L.

Plasma sample preparation, biobanking and biomarker data acquisition and processing: X.Z., M.F.F., M.N.N., D.S.S., M.I.K., O.L.L, and T.K.K.

Statistical analysis and preparation of figures: R.S.

Clinical assessments, coordination and data acquisition: B.E.S., O.L.L

Neuroimaging and neuropathological assessments, data acquisition and processing: D.S.S, T.A.P., A.D.C., M.D.I., J.K.K

Manuscript draft and revision: R.S., X.Z., M.F.F., M.N.N., D.S.S, M.I.K., B.E.S., J.K.K., A.D.C., T.A.P., M.D.I., H.Z., O.L.L., T.K.K

---

**Extended Data Fig. 1.**
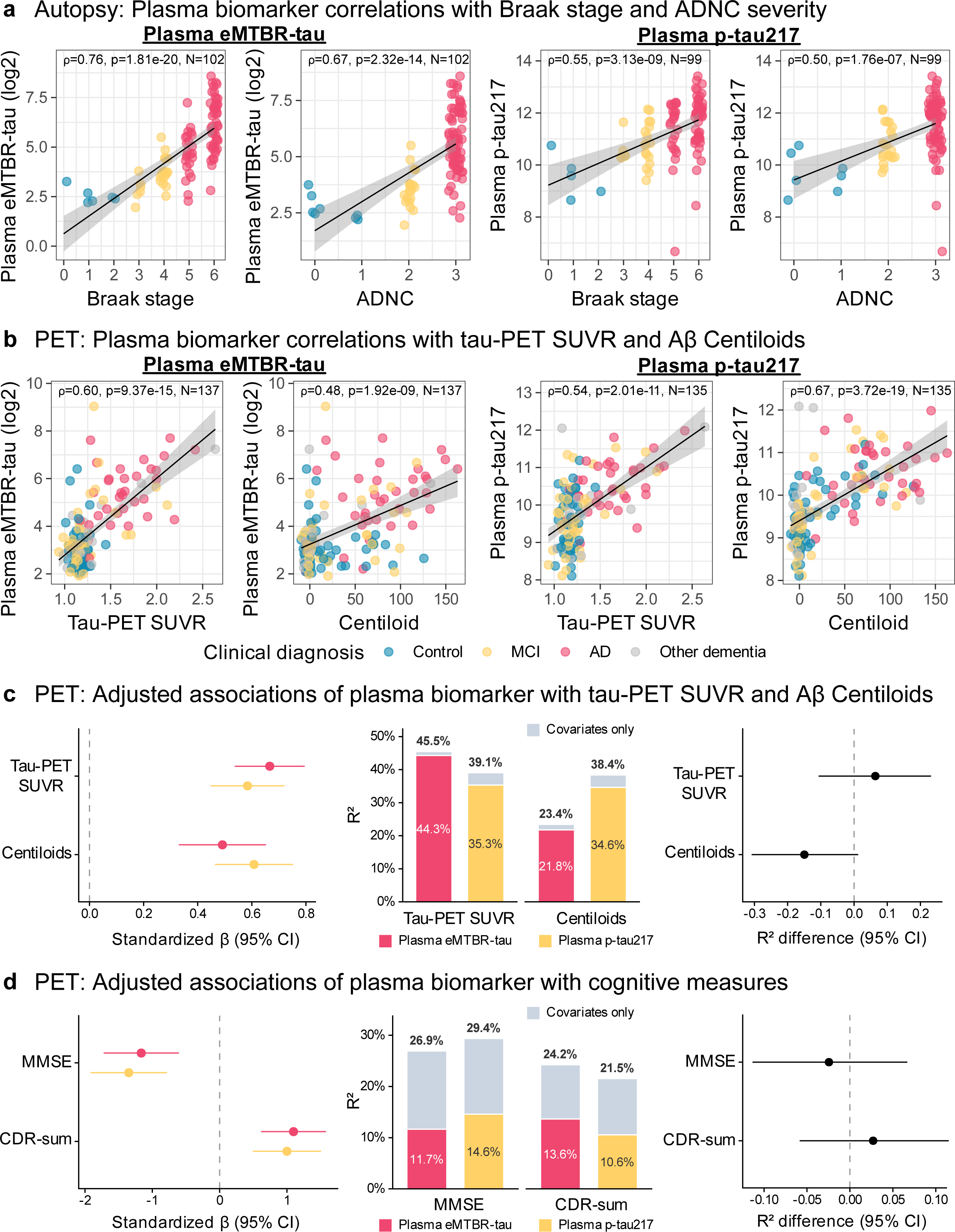
Plasma biomarker associations with neuropathology, PET measures and cognition. **a**, Spearman correlations of plasma eMTBR-tau and p-tau217 with Braak stage and ADNC severity in the autopsy cohort. **b**, Spearman correlations of plasma eMTBR-tau and p-tau217 with tau-PET SUVR and amyloid-PET Centiloid burden in the PET cohort; points are colored by clinical diagnosis. Black lines show fitted linear trends and gray bands show 95% confidence intervals. **c**, Age- and sex-adjusted associations of each plasma biomarker with tau-PET SUVR and Aβ Centiloid burden. Left, coefficients per 1-standard-deviation higher imaging measure with 95% confidence intervals. Center, total model R2 partitioned into biomarker-attributable and covariate-only components. Right, paired-bootstrap differences in model R2 (eMTBR-tau minus p-tau217) with 95% confidence intervals. **d**, Age-, sex-and education-adjusted associations of each plasma biomarker with MMSE and CDR-Sum in the PET cohort, displayed as coefficients with 95% confidence intervals, partitioned model R^2^ and paired-bootstrap differences in model R^2^. Dashed vertical lines mark zero. Abbreviations: AD, Alzheimer disease dementia; MCI, mild cognitive impairment; CI, confidence interval; CDR-Sum, CDR Sum of Boxes; MMSE, Mini-Mental State Examination.

**Extended Data Fig. 2.**
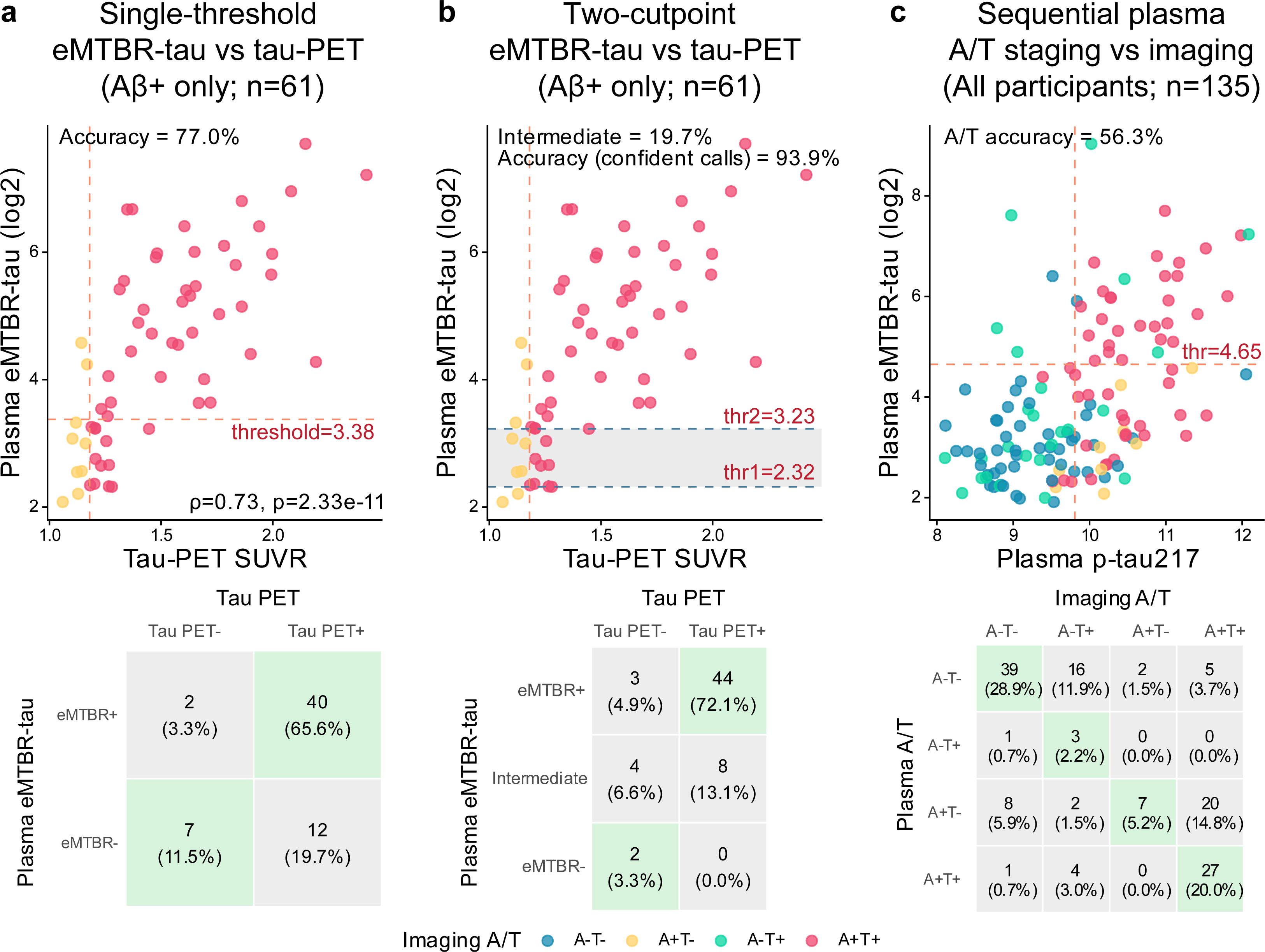
Plasma eMTBR-tau and p-tau217 as surrogates for imaging A/T status. **a**, Single-threshold classification of tau-PET status by plasma eMTBR-tau among amyloid-PET-positive participants. The scatterplot shows the Youden threshold, and the matrix reports counts and percentages for plasma eMTBR-tau and imaging tau-PET classifications. **b**, Two-cutpoint plasma eMTBR-tau classification of tau-PET status among amyloid-PET-positive participants. The shaded region denotes the intermediate zone; accuracy is calculated among confident calls outside this zone. The matrix reports counts and percentages across eMTBR-tau-positive, intermediate and eMTBR-tau-negative classifications. **c**, Sequential plasma A/T staging in all participants. Amyloid status was first classified using the plasma p-tau217 threshold, followed by tau status using the plasma eMTBR-tau threshold among plasma p- tau217-positive participants. The scatterplot shows both thresholds and points colored by imaging A/T group. The matrix cross-tabulates plasma and imaging A/T classifications; diagonal cells are shaded green. Matrix percentages use the panel sample size as denominator. Abbrevations: A, amyloid; T, tau; PET, positron emission tomography; SUVR, standardized uptake value ratio; CDR-Sum, CDR Sum of Boxes; MMSE, Mini-Mental State Examination.

**Extended Data Table 1.**
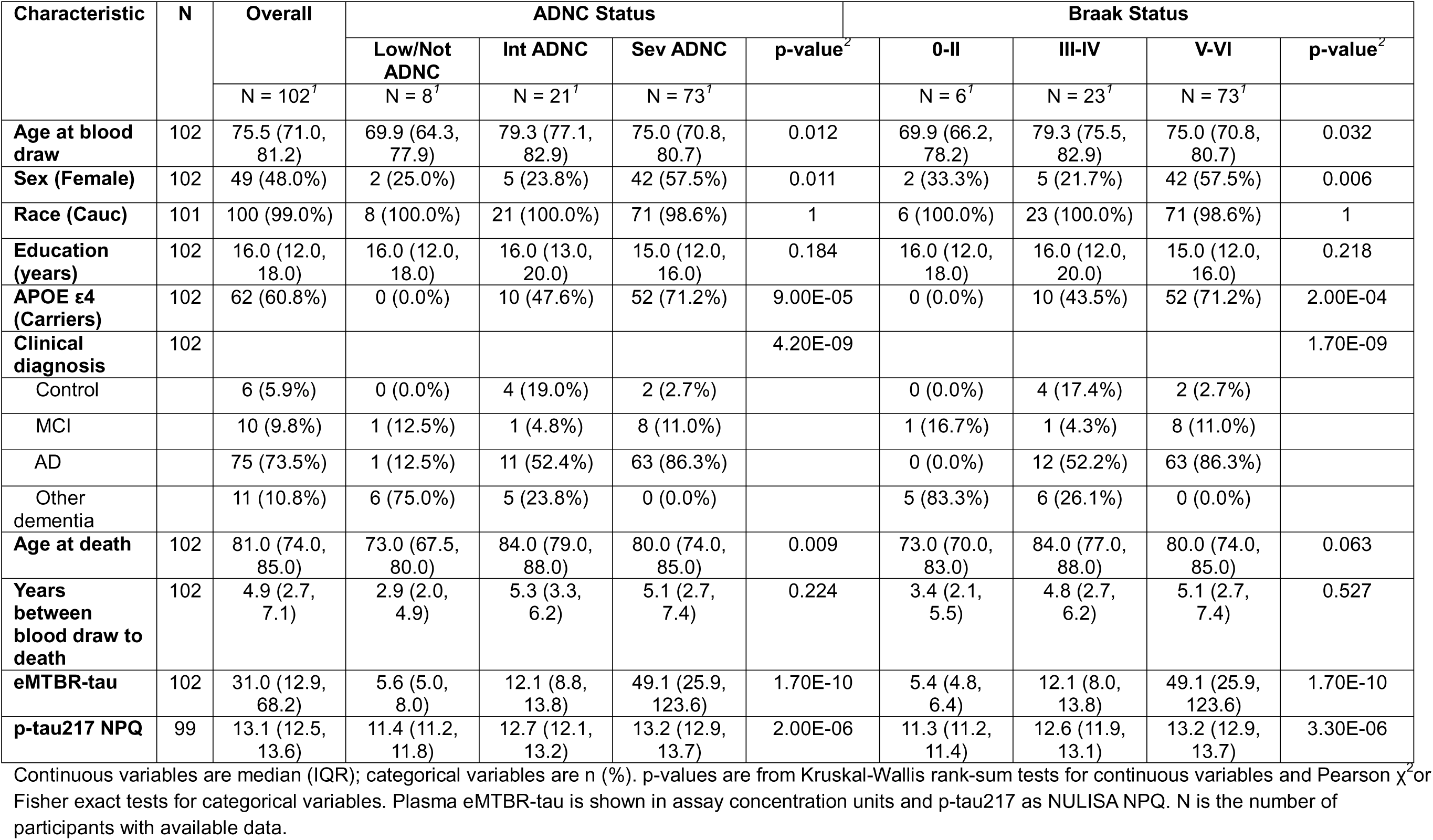
Characteristics of the ADRC-Autopsy cohort by ADNC and Braak stage.

**Extended Data Table 2.** Characteristics of the ADRC-PET cohort by tau PET-defined T status.

| Characteristic | N | Overall | T- | T+ | p-value |
| --- | --- | --- | --- | --- | --- |
|  |  | N = 137 | N = 60 | N = 77 |  |
| **Age at blood draw** | 137 | 73.0 (68.0, 79.0) | 70.5 (66.0, 77.5) | 74.0 (70.0, 81.0) | 0.013 |
| **Sex (Female)** | 137 | 71 (51.8%) | 27 (45.0%) | 44 (57.1%) | 0.158 |
| **Race** | 137 |  |  |  | 0.438 |
| Cauc |  | 119 (86.9%) | 52 (86.7%) | 67 (87.0%) |  |
| BAA |  | 13 (9.5%) | 7 (11.7%) | 6 (7.8%) |  |
| Other/Unknown |  | 5 (3.6%) | 1 (1.7%) | 4 (5.2%) |  |
| **Education (years)** | 136 | 16.0 (14.0, 18.0) | 16.0 (14.0, 18.0) | 16.0 (15.0, 18.0) | 0.147 |
| **APOE ε4 (Carriers)** | 78 | 28 (35.9%) | 11 (28.2%) | 17 (43.6%) | 0.157 |
| **Clinical diagnosis** | 137 |  |  |  | 2.70E-05 |
| Control |  | 45 (32.8%) | 26 (43.3%) | 19 (24.7%) |  |
| MCI |  | 42 (30.7%) | 23 (38.3%) | 19 (24.7%) |  |
| AD |  | 35 (25.5%) | 3 (5.0%) | 32 (41.6%) |  |
| Other dementia |  | 15 (10.9%) | 8 (13.3%) | 7 (9.1%) |  |
| **CDR-Global** | 137 |  |  |  | 0.15 |
| 0 |  | 18 (13.1%) | 11 (18.3%) | 7 (9.1%) |  |
| 0.5 |  | 90 (65.7%) | 41 (68.3%) | 49 (63.6%) |  |
| 1 |  | 22 (16.1%) | 6 (10.0%) | 16 (20.8%) |  |
| 2 |  | 7 (5.1%) | 2 (3.3%) | 5 (6.5%) |  |
| **CDR-Sum** | 135 | 1.5 (0.5, 4.5) | 1.5 (0.5, 2.0) | 3.5 (1.0, 5.0) | 6.90E-04 |
| **MMSE** | 137 | 27.0 (23.0, 29.0) | 27.0 (25.0, 29.0) | 26.0 (23.0, 28.0) | 0.02 |
| **MoCA** | 37 | 23.0 (19.0, 27.0) | 26.0 (21.0, 27.0) | 21.5 (14.0, 28.0) | 0.261 |
| **Amyloid-β Centiloids** | 137 | 15.2 (-0.6, 70.2) | 0.6 (-3.2, 12.0) | 61.3 (3.6, 89.4) | 6.20E-09 |
| **Amyloid-β status (A+)** | 137 | 61 (44.5%) | 9 (15.0%) | 52 (67.5%) | 8.30E-10 |
| **Tau-PET SUVR** | 137 | 1.2 (1.1, 1.4) | 1.1 (1.1, 1.2) | 1.3 (1.2, 1.7) | 1.30E-23 |
| **A/T status** | 137 |  |  |  | 2.40E-40 |
| A-T- |  | 51 (37.2%) | 51 (85.0%) | 0 (0.0%) |  |
| A-T+ |  | 25 (18.2%) | 0 (0.0%) | 25 (32.5%) |  |
| A+T- |  | 9 (6.6%) | 9 (15.0%) | 0 (0.0%) |  |
| A+T+ |  | 52 (38.0%) | 0 (0.0%) | 52 (67.5%) |  |
| **eMTBR-tau** | 137 | 10.0 (6.3, 26.7) | 7.7 (5.6, 10.8) | 19.4 (8.3, 46.8) | 2.50E-07 |
| **p-tau217 NPQ** | 135 | 9.8 (9.1, 10.4) | 9.5 (8.8, 10.0) | 10.2 (9.6, 10.9) | 1.10E-06 |

Continuous variables are median (IQR); categorical variables are n (%). p-values are from Wilcoxon rank-sum tests for continuous variables and Pearson $\chi^2$ or Fisher exact tests for categorical variables. Plasma eMTBR-tau is shown in assay concentration units and p-tau217 as NULISA NPQ. N is the number of participants with available data.
Abbreviations: BAA, Black/African American; Cauc, Caucasian; MoCA, Montreal Cognitive Assessment. T+, tau PET SUVR $\geq 1.18$ ; A+, amyloid PET $\geq 20$ Centiloids.

**Extended Data Table 3.** Performance of plasma eMTBR-tau and p-tau217 for identifying Braak stages V/VI across cumulative blood-draw-to-death intervals in the ADRC-Autopsy cohort. 3a. Sensitivity, specificity and accuracy for Braak stages V/VI vs 0-IV among participants whose blood draw was within 2, 4, 6, 8 or 10 years of death Each marker’s Youden threshold, derived in its full available autopsy sample (eMTBR-tau, log2 concentration; p-tau217, NULISA NPQ), was applied to every window; 95% CIs are exact binomial. 3b. AUCs (DeLong 95% CIs) within each window and paired DeLong comparisons, in participants with both markers.

| Window years | N | N_positive | Threshold | Sensitivity | Sensitivity_lower | Sensitivity_upper | Specificity | Specificity_lower | Specificity_upper | Accuracy | Accuracy_lower | Accuracy_upper |
| --- | --- | --- | --- | --- | --- | --- | --- | --- | --- | --- | --- | --- |
| **<u>eMTBR-tau</u>** |  |  |  |  |  |  |  |  |  |  |  |  |
| 2 | 17 | 12 | 4.45 | 1.00 | 0.74 | 1.00 | 0.80 | 0.28 | 0.99 | 0.94 | 0.71 | 1.00 |
| 4 | 43 | 30 | 4.45 | 0.93 | 0.78 | 0.99 | 0.85 | 0.55 | 0.98 | 0.91 | 0.78 | 0.97 |
| 6 | 63 | 44 | 4.45 | 0.91 | 0.78 | 0.97 | 0.89 | 0.67 | 0.99 | 0.90 | 0.80 | 0.96 |
| 8 | 85 | 59 | 4.45 | 0.92 | 0.81 | 0.97 | 0.92 | 0.75 | 0.99 | 0.92 | 0.84 | 0.97 |
| 10 | 102 | 73 | 4.45 | 0.82 | 0.71 | 0.90 | 0.93 | 0.77 | 0.99 | 0.85 | 0.77 | 0.92 |
| **<u>p-tau217 NPQ</u>** |  |  |  |  |  |  |  |  |  |  |  |  |
| 2 | 17 | 12 | 12.80 | 0.83 | 0.52 | 0.98 | 0.60 | 0.15 | 0.95 | 0.76 | 0.50 | 0.93 |
| 4 | 40 | 30 | 12.80 | 0.77 | 0.58 | 0.90 | 0.60 | 0.26 | 0.88 | 0.73 | 0.56 | 0.85 |
| 6 | 60 | 44 | 12.80 | 0.82 | 0.67 | 0.92 | 0.56 | 0.30 | 0.80 | 0.75 | 0.62 | 0.85 |
| 8 | 82 | 59 | 12.80 | 0.81 | 0.69 | 0.90 | 0.65 | 0.43 | 0.84 | 0.77 | 0.66 | 0.85 |
| 10 | 99 | 73 | 12.80 | 0.79 | 0.68 | 0.88 | 0.69 | 0.48 | 0.86 | 0.77 | 0.67 | 0.85 |

| Window years | N | N_positive | AUC_eMTBR | AUC_lower | AUC_upper | AUC_pTau217 | AUC_lower | AUC_upper | ΔAUC | Z | P | P.adj |
| --- | --- | --- | --- | --- | --- | --- | --- | --- | --- | --- | --- | --- |
| 2 | 17 | 12 | 0.95 | 0.84 | 1.00 | 0.82 | 0.59 | 1.00 | 0.13 | 1.22 | 0.223 | 0.223 |
| 4 | 40 | 30 | 0.92 | 0.81 | 1.00 | 0.76 | 0.59 | 0.93 | 0.16 | 1.72 | 0.085 | 0.106 |
| 6 | 60 | 44 | 0.93 | 0.85 | 0.99 | 0.79 | 0.67 | 0.91 | 0.14 | 2.22 | 0.027 | 0.044 |
| 8 | 82 | 59 | 0.95 | 0.89 | 0.99 | 0.81 | 0.70 | 0.91 | 0.14 | 2.66 | 0.008 | 0.026 |
| 10 | 99 | 73 | 0.92 | 0.87 | 0.97 | 0.81 | 0.72 | 0.90 | 0.11 | 2.56 | 0.010 | 0.026 |

ΔAUC is AUC(eMTBR-tau) - AUC(p-tau217); P.adj is Benjamini–Hochberg adjusted across windows. N\_positive is the number of Braak V/VI cases.

## Methods

### Study design and participants

We analyzed plasma samples from two University of Pittsburgh Alzheimer’s Disease Research Center cohorts spanning cognitively unimpaired controls, MCI, AD and other dementias. Detailed participant recruitment strategies and study design were described in prior publication.^26^ ADRC-Autopsy included 102 brain donors with antemortem SPEAR eMTBR-tau and postmortem Braak stage and ADNC assessment. ADRC-PET included 137 participants with SPEAR eMTBR-tau, clinical assessment, amyloid PET and tau PET. The earliest eligible plasma eMTBR-tau measurement was used for cross-sectional analyses. Longitudinal cognitive analyses in ADRC-Autopsy used all available assessments from the blood-draw visit onward. Informed consent was obtained from all participants; ethics approval details will be added before submission.

### Amyloid-β pathology and tau pathology assessment

Participants underwent [^11^C]Pittsburgh Compound B (PiB) PET to assess cerebral amyloid-β plaque burden.^27–29^ Amyloid burden was expressed on the MRI-independent Centiloid scale, and participants with >20 Centiloids were classified as amyloid positive.^30^

Tau burden was assessed using [^18^F]AV-1451 PET as previously described.^19,31–33^ A composite standardized uptake value ratio (SUVR) was calculated by normalizing uptake across composite Braak regions I-VI to FreeSurfer-defined cerebellar gray matter uptake. Participants with a composite SUVR >1.18 were classified as tau-PET positive and those with an SUVR ≤1.18 as tau-PET negative.^19^

### Neuropathologic assessment

Neuropathologic assessments were performed according to the National Institute on Aging-Alzheimer’s Association (NIA-AA) guidelines^34,35^, following National Alzheimer’s Coordinating Center (NACC) data collection procedures (versions 1, 7, 9, 10, and 11). Neurofibrillary degeneration was classified by Braak stage and grouped as 0-II, III-IV or V-VI. AD neuropathologic change (ADNC) was categorized as none, low, intermediate or high. The primary binary autopsy outcomes were Braak V-VI, high ADNC, and intermediate or high ADNC.

### Plasma p-tau217

Plasma p-tau217 was measured in the Department of Psychiatry at the University of Pittsburgh using the NULISAseq CNS Disease Panel 120 on an ArgoHT system (Alamar Biosciences, Inc.). Experimental procedures have been described previously.^31,36,37^ Briefly, paired capture and detection antibodies carrying target-specific DNA barcodes were used in a double capture-and-release workflow, followed by ligation, sample barcoding and next-generation sequencing. Counts were normalized to internal and inter-plate controls, log2-transformed and reported as NULISA Protein Quantification values. Only p-tau217 was used as a comparator in the present study.

### Plasma eMTBR-tau

Plasma eMTBR-tau was quantified using the singleplex SPEAR TrueDetect^TM^ eMTBR-Tau immunoassay (Spear Bio, Woburn, Massachusetts) following the vendor’s instructions. The assay uses proprietary Successive Proximity Extension Amplification Reaction technology, a homogeneous, wash-free immunoassay workflow with quantitative polymerase chain reaction readout and requires 1 μL of diluted plasma. Concentrations were reported in pg/mL (log2), and values below the functional lower limit of detection were treated as missing.

### Statistical analysis

All statistical analyses were performed using R version 4.5.1 (R Foundation for Statistical Computing, Vienna, Austria). Continuous variables were summarized as medians with interquartile ranges (IQRs), and categorical variables as counts and percentages. Two-group comparisons of continuous variables used Wilcoxon rank-sum tests. Comparisons across more than two groups used Kruskal-Wallis tests, followed by Dunn pairwise tests with Bonferroni correction. Fold changes were calculated as ratios of group means. Categorical variables were compared using Pearson χ^2^ or Fisher exact tests, as appropriate. All tests were two-sided. Analyses used available complete cases without imputation. Each marker was analyzed in all participants with available measurements, while head-to-head comparisons between eMTBR-tau and p-tau217 were restricted to participants with both markers.

Spearman rank correlations quantified associations with continuous imaging and neuropathologic measures, and differences between markers were tested by paired bootstrap (5,000 resamples). Linear regressions were conducted between z-scale standardized markers and imaging measures (adjusted for age at blood draw and sex) and cognitive measures (adjusted for age at blood draw, sex and education years). Partial R^2^ was calculated from its t statistic and residual degrees of freedom. Marker differences in model R² were tested by paired bootstrap (1,000 resamples).

Receiver operating characteristic (ROC) analyses evaluated discrimination of amyloid-PET positivity, tau-PET positivity, tau-PET positivity among Aβ+ participants and combined A+T+ status in ADRC-PET, and Braak stages V/VI and high ADNC in ADRC-Autopsy. AUC 95% CIs used the DeLong method; Youden thresholds defined sensitivity, specificity, predictive values and accuracy; correlated AUCs were compared by paired DeLong tests.

For A/T classification (amyloid PET ≥20 Centiloids; tau-PET SUVR ≥1.18), eMTBR-tau thresholds for tau PET status among Aβ+ participants were derived by Youden index and by a two-cutpoint approach maximizing accuracy among confidently classified participants, with an intermediate zone <20%. Sequential staging applied a p-tau217 Youden threshold for amyloid status, then an eMTBR-tau threshold derived among p-tau217-positive participants and was also evaluated after excluding A-T+ participants.

Longitudinal MMSE and CDR-Sum in ADRC-Autopsy were modeled by linear mixed-effects models with time, the standardized baseline marker and their interaction, adjusted for age at blood draw, sex and education, with random intercepts and slopes (random intercepts only if singular) and Satterthwaite degrees of freedom. Time-varying models related cognition to the nearest preceding blood draw, separating between- and within-person effects by person-mean centering. Markers were compared in joint models by Wald tests of coefficient differences.

Temporal robustness was assessed within cumulative blood-draw-to-death intervals (2-10 years), applying each marker’s full-sample Youden threshold to estimate AUCs (DeLong CIs) and operating characteristics (exact binomial CIs); AUCs were compared by paired DeLong tests with BH correction across intervals.

## Code availability

No specialized code was employed in the preparation of this manuscript.

## Ethics/IRB statement

The Pitt ADRC study was approved by the University of Pittsburgh Institutional Review Board (STUDY19110245), and all participants provided written informed consent. Neuropathological assessment of donor brains was approved by the Committee for Oversight of Research and Clinical Training Involving Decedents (CORID) and the appropriate institutional review boards at the University of Pittsburgh, in accordance with the Declaration of Helsinki.

