## Supplementary Tables for "Plasma eMTBR-tau as a robust biomarker for Alzheimer’s disease tangle pathology up to 10 years before death"

**Supplementary Table 1.** Pairwise comparisons of plasma eMTBR-tau across clinical and pathologic groups.

1a. ADRC-Autopsy cohort, by clinical diagnosis, ADNC, Braak stage, Thal phase and CERAD score (CERAD C1 excluded because of no subjects was classified in this group).

| **group** | **group1** | **group2** | **n1** | **n2** | **fc** | **p** | **p.adj** |
| --- | --- | --- | --- | --- | --- | --- | --- |
| Clinical diagnosis | Control | MCI | 6 | 10 | 2.14 | 0.118 | 0.142 |
| Clinical diagnosis | Control | AD | 6 | 75 | 4.50 | 0.003 | 0.007 |
| Clinical diagnosis | Control | Other dementia | 6 | 11 | 0.72 | 0.591 | 0.591 |
| Clinical diagnosis | MCI | AD | 10 | 75 | 2.11 | 0.046 | 0.069 |
| Clinical diagnosis | MCI | Other dementia | 10 | 11 | 0.34 | 0.004 | 0.007 |
| Clinical diagnosis | AD | Other dementia | 75 | 11 | 0.16 | 1.09E-06 | 6.54E-06 |
| ADNC | Low/Not ADNC | Int ADNC | 8 | 21 | 1.88 | 0.005 | 0.005 |
| ADNC | Low/Not ADNC | Sev ADNC | 8 | 73 | 8.08 | 1.20E-05 | 1.80E-05 |
| ADNC | Int ADNC | Sev ADNC | 21 | 73 | 4.29 | 2.92E-08 | 8.76E-08 |
| Braak stage | 0-II | III-IV | 6 | 23 | 2.03 | 0.002 | 0.002 |
| Braak stage | 0-II | V-VI | 6 | 73 | 8.95 | 9.80E-05 | 1.47E-04 |
| Braak stage | III-IV | V-VI | 23 | 73 | 4.41 | 6.20E-09 | 1.86E-08 |
| Thal phase | A0 | A1 | 5 | 4 | 0.60 | 0.016 | 0.024 |
| Thal phase | A0 | A2 | 5 | 3 | 0.99 | 1 | 1 |
| Thal phase | A0 | A3 | 5 | 70 | 5.09 | 0.001 | 0.003 |
| Thal phase | A1 | A2 | 4 | 3 | 1.64 | 0.057 | 0.069 |
| Thal phase | A1 | A3 | 4 | 70 | 8.43 | 0.001 | 0.003 |
| Thal phase | A2 | A3 | 3 | 70 | 5.14 | 0.01 | 0.02 |
| CERAD score | C0 | C2 | 8 | 11 | 1.78 | 0.026 | 0.026 |
| CERAD score | C0 | C3 | 8 | 83 | 6.83 | 1.85E-05 | 5.55E-05 |
| CERAD score | C2 | C3 | 11 | 83 | 3.85 | 8.54E-05 | 1.28E-04 |

1b. ADRC-PET cohort, by clinical diagnosis, clinical diagnosis-by-Aβ status, Aβ status, T status and A/T status.

| **group** | **group1** | **group2** | **n1** | **n2** | **fc** | **p** | **p.adj** |
| --- | --- | --- | --- | --- | --- | --- | --- |
| Clinical diagnosis | Control | MCI | 45 | 42 | 1.42 | 0.049 | 0.073 |
| Clinical diagnosis | Control | AD | 45 | 35 | 4.47 | 1.86E-10 | 1.12E-09 |
| Clinical diagnosis | Control | Other dementia | 45 | 15 | 1.54 | 0.143 | 0.172 |
| Clinical diagnosis | MCI | AD | 42 | 35 | 3.15 | 6.48E-07 | 1.94E-06 |
| Clinical diagnosis | MCI | Other dementia | 42 | 15 | 1.08 | 0.795 | 0.795 |
| Clinical diagnosis | AD | Other dementia | 35 | 15 | 0.34 | 9.67E-04 | 0.002 |
| Clinical diagnosis × Aβ status | Control A- | Control A+ | 31 | 14 | 0.83 | 0.799 | 0.83 |
| Clinical diagnosis × Aβ status | Control A- | MCI A- | 31 | 29 | 1.13 | 0.49 | 0.606 |
| Clinical diagnosis × Aβ status | Control A- | MCI A+ | 31 | 13 | 1.96 | 0.013 | 0.033 |
| Clinical diagnosis × Aβ status | Control A- | AD A+ | 31 | 32 | 4.18 | 1.27E-08 | 1.90E-07 |
| Clinical diagnosis × Aβ status | Control A- | Other dementia A- | 31 | 13 | 1.21 | 0.525 | 0.606 |
| Clinical diagnosis × Aβ status | Control A+ | MCI A- | 14 | 29 | 1.36 | 0.376 | 0.513 |
| Clinical diagnosis × Aβ status | Control A+ | MCI A+ | 14 | 13 | 2.37 | 0.008 | 0.023 |
| Clinical diagnosis × Aβ status | Control A+ | AD A+ | 14 | 32 | 5.05 | 5.88E-08 | 2.94E-07 |
| Clinical diagnosis × Aβ status | Control A+ | Other dementia A- | 14 | 13 | 1.47 | 0.325 | 0.488 |
| Clinical diagnosis × Aβ status | MCI A- | MCI A+ | 29 | 13 | 1.74 | 0.028 | 0.053 |
| Clinical diagnosis × Aβ status | MCI A- | AD A+ | 29 | 32 | 3.71 | 4.55E-08 | 2.94E-07 |
| Clinical diagnosis × Aβ status | MCI A- | Other dementia A- | 29 | 13 | 1.08 | 0.83 | 0.83 |
| Clinical diagnosis × Aβ status | MCI A+ | AD A+ | 13 | 32 | 2.13 | 0.022 | 0.047 |
| Clinical diagnosis × Aβ status | MCI A+ | Other dementia A- | 13 | 13 | 0.62 | 0.169 | 0.282 |
| Clinical diagnosis × Aβ status | AD A+ | Other dementia A- | 32 | 13 | 0.29 | 1.31E-04 | 4.91E-04 |
| Aβ status | A- | A+ | 76 | 61 | 2.14 | 4.75E-06 | 4.75E-06 |
| T status | T- | T+ | 60 | 77 | 2.58 | 2.52E-07 | 2.52E-07 |
| A/T status | A-T- | A+T- | 51 | 9 | 0.99 | 0.967 | 0.967 |
| A/T status | A-T- | A-T+ | 51 | 25 | 1.80 | 0.109 | 0.164 |
| A/T status | A-T- | A+T+ | 51 | 52 | 3.06 | 1.28E-08 | 7.68E-08 |
| A/T status | A+T- | A-T+ | 9 | 25 | 1.81 | 0.316 | 0.379 |
| A/T status | A+T- | A+T+ | 9 | 52 | 3.08 | 0.002 | 0.005 |
| A/T status | A-T+ | A+T+ | 25 | 52 | 1.70 | 0.018 | 0.036 |

Groups were compared with two-sided Wilcoxon rank-sum tests; p.adj is Benjamini-Hochberg adjusted within each grouping (unadjusted for two-group comparisons). Fold change (fc) is the ratio of geometric means, group2/group1 (2^difference in mean log2-eMTBR-tau). Abbreviations: Aβ+, amyloid PET ≥20 Centiloids; T+; tau -PET SUVR ≥1.18; ADNC, Alzheimer's disease neuropathologic change; MCI, mild cognitive impairment; AD, Alzheimer's disease dementia.

**Supplementary Table 2.** Associations of plasma eMTBR-tau and p-tau217 with amyloid and tau PET in the ADRC-PET cohort.

| **Marker** | **N** | **β** | **SE** | **CI_low** | **CI_high** | **P** | **Partial_R2** | **Model_R2** | **Adjusted_R2** |
| --- | --- | --- | --- | --- | --- | --- | --- | --- | --- |
| Tau-PET SUVR | | | | | | | | | |
| Plasma eMTBR-tau | 135 | 0.67 | 0.07 | 0.54 | 0.80 | 2.38E-18 | 44.3% | 45.5% | 44.2% |
| Plasma p-tau217 | 135 | 0.58 | 0.07 | 0.45 | 0.72 | 4.58E-14 | 35.3% | 39.1% | 37.7% |
| Centiloids | | | | | | | | | |
| Plasma eMTBR-tau | 135 | 0.49 | 0.08 | 0.33 | 0.65 | 1.51E-08 | 21.8% | 23.4% | 21.7% |
| Plasma p-tau217 | 135 | 0.61 | 0.07 | 0.46 | 0.75 | 9.38E-14 | 34.6% | 38.4% | 37.0% |

Linear regression with the standardized plasma marker as the outcome and the standardized PET measure (tau-PET SUVR or Centiloids) as the predictor, adjusted for age at blood draw and sex, in participants with both plasma markers (n=135). Abbreviations: β, SD change in plasma marker per 1-SD higher PET measure, with 95% CI; Partial R^2^, the variance explained by the PET term, calculated from its t statistic and residual degrees of freedom; Model R^2^ and adjusted R^2^, the full model including covariates.

**Supplementary Table 3.** Diagnostic performance of plasma eMTBR-tau and p-tau217.

3a. ADRC-Autopsy cohort: discrimination of Braak stages V/VI vs 0–IV (first two rows) and high ADNC vs not high (last two rows), in each marker's available sample.

| **Marker** | **AUC** | **Lower** | **Upper** | **Threshold** | **Sensitivity** | **Specificity** | **PPV** | **NPV** | **Accuracy** |
| --- | --- | --- | --- | --- | --- | --- | --- | --- | --- |
| ***Braak V-VI vs 0-IV*** | | | | | | | | | |
| eMTBR-tau | 0.92 | 0.87 | 0.98 | 4.45 | 0.82 | 0.96 | 0.98 | 0.66 | 0.86 |
| p-tau217 | 0.81 | 0.72 | 0.90 | 12.80 | 0.79 | 0.69 | 0.88 | 0.55 | 0.77 |
| ***High ADNC vs. Not High*** | | | | | | | | | |
| eMTBR-tau | 0.92 | 0.87 | 0.98 | 4.45 | 0.82 | 0.96 | 0.98 | 0.66 | 0.86 |
| p-tau217 | 0.81 | 0.72 | 0.90 | 12.80 | 0.79 | 0.69 | 0.88 | 0.55 | 0.77 |

3b. Paired DeLong comparisons of the AUCs in (a), in participants with both markers.

| **Task** | **Comparison** | **N** | **N_cases** | **AUC_eMTBR** | **AUC_pTau217** | **ΔAUC** | **Z** | **P** |
| --- | --- | --- | --- | --- | --- | --- | --- | --- |
| Braak V/VI vs 0-IV | eMTBR-tau vs p-tau217 | 99 | 73 | 0.92 | 0.81 | 0.11 | 2.56 | 0.01 |
| ADNC High vs Not High | eMTBR-tau vs p-tau217 | 99 | 73 | 0.92 | 0.81 | 0.11 | 2.56 | 0.01 |

3c. ADRC-PET cohort: discrimination of T+ vs T-, A+ vs A−, T+ vs T- among A+ participants, and A+T+ vs all others.

| **Task** | **Marker** | **AUC** | **Lower** | **Upper** | **Threshold** | **Sensitivity** | **Specificity** | **PPV** | **NPV** | **Accuracy** |
| --- | --- | --- | --- | --- | --- | --- | --- | --- | --- | --- |
| T+ vs T- | eMTBR-tau | 0.75 | 0.67 | 0.83 | 3.59 | 0.64 | 0.79 | 0.80 | 0.62 | 0.70 |
| T+ vs T- | p-tau217 | 0.75 | 0.66 | 0.83 | 9.84 | 0.68 | 0.74 | 0.78 | 0.63 | 0.70 |
| A+ vs A- | eMTBR-tau | 0.72 | 0.63 | 0.81 | 3.94 | 0.61 | 0.81 | 0.73 | 0.71 | 0.72 |
| A+ vs A- | p-tau217 | 0.90 | 0.84 | 0.95 | 9.81 | 0.89 | 0.80 | 0.78 | 0.89 | 0.84 |
| T+ vs T- within A+ | eMTBR-tau | 0.83 | 0.70 | 0.96 | 3.38 | 0.77 | 0.78 | 0.95 | 0.37 | 0.77 |
| T+ vs T- within A+ | p-tau217 | 0.62 | 0.42 | 0.82 | 10.64 | 0.42 | 0.89 | 0.96 | 0.21 | 0.49 |
| A+T+ vs others | eMTBR-tau | 0.77 | 0.69 | 0.86 | 3.94 | 0.67 | 0.81 | 0.69 | 0.80 | 0.76 |
| A+T+ vs others | p-tau217 | 0.87 | 0.81 | 0.93 | 9.73 | 0.94 | 0.70 | 0.66 | 0.95 | 0.79 |

3d. Paired DeLong comparisons of the AUCs in (c)

| **Task** | **Comparison** | **N** | **N_cases** | **AUC_eMTBR** | **AUC_pTau217** | **ΔAUC** | **Z** | **P** |
| --- | --- | --- | --- | --- | --- | --- | --- | --- |
| T+ vs T- | eMTBR-tau vs p-tau217 | 135 | 77 | 0.750 | 0.746 | 0.004 | 0.083 | 0.934 |
| A+ vs A- | eMTBR-tau vs p-tau217 | 135 | 61 | 0.721 | 0.897 | -0.176 | -4.008 | 6.13E-05 |
| T+ vs T- within A+ | eMTBR-tau vs p-tau217 | 61 | 52 | 0.829 | 0.622 | 0.207 | 2.987 | 0.003 |
| A+T+ vs others | eMTBR-tau vs p-tau217 | 135 | 52 | 0.774 | 0.872 | -0.098 | -2.276 | 0.023 |

AUC 95% CIs were estimated by the DeLong method. Thresholds are Youden-index optimal cut-offs (eMTBR-tau, log2 concentration; p-tau217, NULISA NPQ), at which sensitivity, specificity, PPV, NPV and accuracy are reported. Abbreviations: ΔAUC, AUC(eMTBR-tau) - AUC(p-tau217); N_cases. the number of positive cases; PPV/NPV, positive/negative predictive value.

**Supplementary Table 4.** Associations of plasma eMTBR-tau and p-tau217 with cross-sectional and longitudinal cognition in the ADRC-Autopsy cohort.

4a. Cross-sectional linear regression of MMSE or CDR-Sum on each plasma marker at blood draw.

| **Outcome** | **N** | **β** | **SE** | **CI_low** | **CI_high** | **t_value** | **DF** | **P** | **Partial_R2** | **Model_R2** | **Adjusted_R2** |
| --- | --- | --- | --- | --- | --- | --- | --- | --- | --- | --- | --- |
| ***eMTBR-tau*** | | | | | | | | | | | |
| MMSE | 101 | -3.34 | 0.65 | -4.63 | -2.05 | -5.15 | 96 | 1.38E-06 | 21.7% | 27.5% | 24.5% |
| CDR-sum | 92 | 1.83 | 0.39 | 1.05 | 2.61 | 4.66 | 87 | 1.13E-05 | 20.0% | 25.9% | 22.5% |
| ***p-tau217 NPQ*** | | | | | | | | | | | |
| MMSE | 98 | -1.00 | 0.70 | -2.38 | 0.38 | -1.44 | 93 | 0.154 | 2.2% | 8.0% | 4.1% |
| CDR-sum | 90 | 0.80 | 0.42 | -0.02 | 1.63 | 1.93 | 85 | 0.056 | 4.2% | 10.2% | 5.9% |

β is the change in score per 1-SD higher marker, with 95% CI and partial R².

4b. Linear mixed-effects models comparing annual rates of cognitive change between participants above vs below each marker's Youden threshold for Braak stages V/VI.

| **Outcome** | **n_subjects** | **n_obs** | **model_used** | **estimate** | **SE** | **statistic** | **df** | **P** |
| --- | --- | --- | --- | --- | --- | --- | --- | --- |
| ***eMTBR-tau*** | | | | | | | | |
| MMSE | 101 | 347 | full (random slope) | -1.08 | 0.42 | -2.58 | 42.43 | 0.01 |
| CDR-Sum | 92 | 338 | full (random slope) | 0.58 | 0.28 | 2.09 | 58.01 | 0.04 |
| ***p-tau217 NPQ*** | | | | | | | | |
| MMSE | 98 | 342 | full (random slope) | -0.81 | 0.45 | -1.81 | 42.90 | 0.08 |
| CDR-Sum | 90 | 332 | full (random slope) | 0.50 | 0.30 | 1.66 | 56.67 | 0.10 |

4c. Linear mixed-effects models of the continuous baseline marker-by-time interaction: additional annual change per 1-SD higher baseline marker.

| **Outcome** | **n_subjects** | **n_obs** | **model_used** | **effect** | **term** | **estimate** | **std.error** | **statistic** | **df** | **p.value** |
| --- | --- | --- | --- | --- | --- | --- | --- | --- | --- | --- |
| ***eMTBR-tau*** | | | | | | | | | | |
| MMSE | 101 | 347 | full (random slope) | fixed | z_emtbr_bl:years_from_draw | -0.76 | 0.23 | -3.30 | 46.00 | 0.002 |
| CDR-Sum | 92 | 338 | full (random slope) | fixed | z_emtbr_bl:years_from_draw | 0.36 | 0.16 | 2.30 | 65.12 | 0.02 |
| ***p-tau217 NPQ*** | | | | | | | | | | |
| MMSE | 98 | 342 | full (random slope) | fixed | z_ptau_bl:years_from_draw | -0.38 | 0.22 | -1.75 | 38.26 | 0.09 |
| CDR-Sum | 90 | 332 | full (random slope) | fixed | z_ptau_bl:years_from_draw | 0.21 | 0.15 | 1.40 | 54.14 | 0.17 |

The estimate is the group-by-time interaction, indicating the difference in annual change (points/year), High - Low.

4d. Time-varying models relating each cognitive assessment to the marker value at the nearest preceding blood draw

| **Outcome** | **n_subjects** | **n_obs** | **estimate** | **std.error** | **statistic** | **df** | **p.value** |
| --- | --- | --- | --- | --- | --- | --- | --- |
| ***eMTBR-tau*** | | | | | | | |
| MMSE | 101 | 347 | -3.49 | 0.54 | -6.42 | 164.13 | 1.43E-09 |
| CDR-Sum | 92 | 338 | 2.37 | 0.34 | 6.98 | 187.59 | 4.97E-11 |
| ***p-tau217 NPQ*** | | | | | | | |
| MMSE | 99 | 448 | -0.71 | 0.41 | -1.75 | 346.50 | 0.08 |
| CDR-Sum | 94 | 437 | -0.03 | 0.25 | -0.11 | 387.95 | 0.91 |

4e. The time-varying association in (d) split into within-person (change from the person's mean) and between-person (person mean) components by person-mean centering.

| **Outcome** | **n_subjects** | **n_obs** | **effect** | **term** | **estimate** | **std.error** | **statistic** | **df** | **p.value** |
| --- | --- | --- | --- | --- | --- | --- | --- | --- | --- |
| ***eMTBR-tau*** | | | | | | | | | |
| MMSE | 101 | 347 | fixed | z_emtbr_within | -2.87 | 1.11 | -2.58 | 181.03 | 0.01 |
| MMSE | 101 | 347 | fixed | z_emtbr_between | -3.71 | 0.64 | -5.76 | 89.35 | 1.17E-07 |
| CDR-Sum | 92 | 338 | fixed | z_emtbr_within | 2.76 | 0.60 | 4.62 | 197.10 | 6.78E-06 |
| CDR-Sum | 92 | 338 | fixed | z_emtbr_between | 2.16 | 0.43 | 5.06 | 86.14 | 2.38E-06 |
| ***p-tau217 NPQ*** | | | | | | | | | |
| MMSE | 99 | 448 | fixed | z_ptau_within | -0.03 | 0.50 | -0.07 | 246.20 | 0.95 |
| MMSE | 99 | 448 | fixed | z_ptau_between | -2.01 | 0.65 | -3.06 | 49.60 | 3.52E-03 |
| CDR-Sum | 94 | 437 | fixed | z_ptau_within | -0.40 | 0.29 | -1.41 | 283.44 | 0.16 |
| CDR-Sum | 94 | 437 | fixed | z_ptau_between | 1.01 | 0.44 | 2.27 | 67.72 | 0.03 |

4f. Head-to-head joint models including both markers in participants with both markers

| **Outcome** | **Contrast** | **N_subjects** | **N_obs** | **β_1** | **SE_1** | **P_1** | **β_2** | **SE_2** | **P_2** | **Δβ** | **ΔSE** | **Z** | **P_value** |
| --- | --- | --- | --- | --- | --- | --- | --- | --- | --- | --- | --- | --- | --- |
| ***Baseline value (continuous) x time*** | | | | | | | | | | | | | |
| MMSE | eMTBR-tau - p-tau217 | 98 | 342 | -0.01 | 0.25 | 0.98 | -0.74 | 0.29 | 0.01 | -0.73 | 0.48 | -1.53 | 0.13 |
| CDR-Sum | eMTBR-tau - p-tau217 | 90 | 332 | -0.01 | 0.19 | 0.98 | 0.36 | 0.21 | 0.09 | 0.36 | 0.37 | 0.99 | 0.32 |
| ***Baseline Youden group (High vs Low) x time*** | | | | | | | | | | | | | |
| MMSE | eMTBR-tau - p-tau217 | 98 | 342 | -0.46 | 0.47 | 0.33 | -0.87 | 0.45 | 0.06 | -0.40 | 0.77 | -0.53 | 0.60 |
| CDR-Sum | eMTBR-tau - p-tau217 | 90 | 332 | 0.30 | 0.33 | 0.36 | 0.45 | 0.31 | 0.15 | 0.15 | 0.53 | 0.28 | 0.78 |
| ***Time-varying current value (pooled)*** | | | | | | | | | | | | | |
| MMSE | eMTBR-tau - p-tau217 | 98 | 342 | 1.06 | 0.45 | 0.02 | -3.94 | 0.58 | 9.25E-11 | -5.00 | 0.84 | -5.93 | 3.12E-09 |
| CDR-Sum | eMTBR-tau - p-tau217 | 90 | 332 | -0.75 | 0.26 | 0.00 | 2.65 | 0.35 | 7.51E-13 | 3.40 | 0.48 | 7.03 | 2.03E-12 |

Beta_1 (p-tau217) and Beta_2 (eMTBR-tau) are mutually adjusted, and Beta_diff (Beta_2 − Beta_1) was tested with a Wald z test based on the fixed-effect variance-covariance matrix. Mixed models (**b-f**) included participant-specific random intercepts and slopes for time, fitting a random intercept only when the random-slope model was singular. Degrees of freedom were estimated by the Satterthwaite approximation. Abbreviations: n_subjects, number of participants; n_obs, number of cognitive assessments; se, standard error; df, degree of freedom; CDR-Sum, CDR Sum of Boxes; MMSE, Mini-Mental State Examination.
